# SVEP1 Upregulation Is a Prominent Molecular Response to Dietary Sodium Loading and Correlates with Inverse Salt Sensitivity

**DOI:** 10.1101/2025.08.12.25333544

**Authors:** Satish RamachandraRao, Christiana Hench, Andrea Berrido, Richard J. Auchus, Jonathan Troost, John Darcy, James Brian Byrd

**Author notes:** Corresponding Author: James Brian Byrd, MD, MS, 5570C Medical Science Research, Building II, 1150 W. Medical Center Drive, Ann Arbor, MI 48109.

## Abstract

**Background:** While some individuals exhibit salt sensitivity, others demonstrate salt resistance or inverse salt sensitivity—blood pressure reduction during high sodium intake. The molecular mechanisms underlying heterogeneous blood pressure responses to dietary sodium remain poorly understood. Deep proteomics provides a new tool to identify molecular mediators of salt resistance and inverse salt sensitivity.

**Methods:** We conducted a randomized crossover trial in 20 normotensive adults comparing 8-day periods of low-sodium (10 mmol/day) versus high-sodium (300 mmol/day) diets. Comprehensive plasma proteomic analysis was performed using SomaLogic’s 7k proteomics platform, which measures approximately 7,000 human proteins. The change in proteins between the high- and low-sodium diets was compared with the change in blood pressure.

**Results:** Despite average weight difference of +1.4 kg during high-versus low-sodium intake (p=8.85×10^-7^), diastolic blood pressure and mean arterial pressure were significantly lower (67.0±7.5 vs 69.7±8.0 mm Hg, p=0.014 for diastolic blood pressure; 82.2±7.6 versus 84.8±8.3 mm Hg, p=0.029 for mean arterial pressure). Among approximately 7,000 proteins analyzed, SVEP1 demonstrated one of the most significant responses to sodium loading, with two independent aptamers (antibody-like DNA molecules) ranking 4th (p= 5.33×10^-6^) and 8th (p=2.19×10^-5^) in statistical significance. SVEP1 substantially outranked established sodium-regulatory hormones including renin (23rd) and NT-proBNP (16th). SVEP1 upregulation correlated inversely with blood pressure changes (R= -0.50, p=0.028), and individuals exhibiting inverse salt sensitivity demonstrated 2-fold higher SVEP1 responses. Changes in SVEP1 correlated strongly with changes in NT-ProBNP (R= 0.80, p<0.001). Reactome analysis revealed coordinated extracellular matrix remodeling as the dominant biological response to sodium loading.

**Conclusions:** SVEP1 emerges as a primary molecular correlate of blood pressure responses to dietary sodium, likely through a volume- or stretch-mediated stimulus. Given SVEP1’s established functions in vascular smooth muscle relaxation and lymphangiogenesis, these findings suggest novel pathways mediating cardiovascular adaptation to sodium challenges and potential biomarkers for identifying salt-sensitive versus salt-resistant individuals.

## Introduction

Over a century ago, Ambard and Beaujard observed that salt loading produced variable blood pressure responses, distinguishing between those who “accommodate themselves” and others who develop “permanent hypertension.”^1^ Despite this early recognition of heterogeneous responses to salt, the molecular mechanisms underlying individual salt sensitivity have remained elusive. While approximately 50% of hypertensive and 25% of normotensive individuals demonstrate salt sensitivity, others show salt resistance.^2-5^ Most intriguingly, a substantial subset exhibits inverse salt sensitivity, experiencing blood pressure reduction during high sodium intake.^6-12^ Traditional models of sodium metabolism poorly explain this wide heterogeneity, which challenges uniform dietary recommendations and highlights the need for precision medicine approaches to sodium management.

The molecular basis for these diverse responses remains largely unknown. A predominant focus on renal mechanisms incompletely explains the vascular adaptations that enable most individuals to maintain normal blood pressure despite documented volume expansion during high sodium intake. Sullivan and Ratts demonstrated that humans vasodilate in response to chronic sodium loading, reducing systemic vascular resistance by 19%.^13^ Failure to vasodilate during sodium loading is associated with increased salt sensitivity in normotensive and borderline hypertensive people.^14^ A failure to vasodilate in response to dietary sodium is also a factor in salt sensitivity in mice.^15^ The molecular mediators of this protective vascular response, however, have remained elusive for four decades.

Recent advances in proteomics permit the measurement of nearly 7,000 proteins in a small volume of plasma. We hypothesized that unbiased proteomic analysis at this depth during controlled sodium loading would reveal novel molecular pathways underlying heterogeneous sodium responses. The SomaLogic SOMAscan 7k platform, which uses single-stranded DNA aptamers to quantify thousands of proteins simultaneously, provided an opportunity to identify sodium-responsive proteins without *a priori* mechanistic assumptions.

## Methods

### Study Design and Participants

We conducted a randomized crossover trial (Sodium Diet Effect on Aldosterone and Urinary RNA [SALTY]; NCT04168073; **Figure 1**) in 20 normotensive adults aged 21-50 years with blood pressure less than 140/90 mmHg who had never been prescribed antihypertensive medications. The SALTY trial excluded participants with cardiovascular disease, kidney disease, diabetes, pregnancy, or conditions that might compromise safety or study integrity. The study was approved by the University of Michigan Institutional Review Board (protocol HUM00097366) and conducted in accordance with the Declaration of Helsinki.

**Figure 1.**
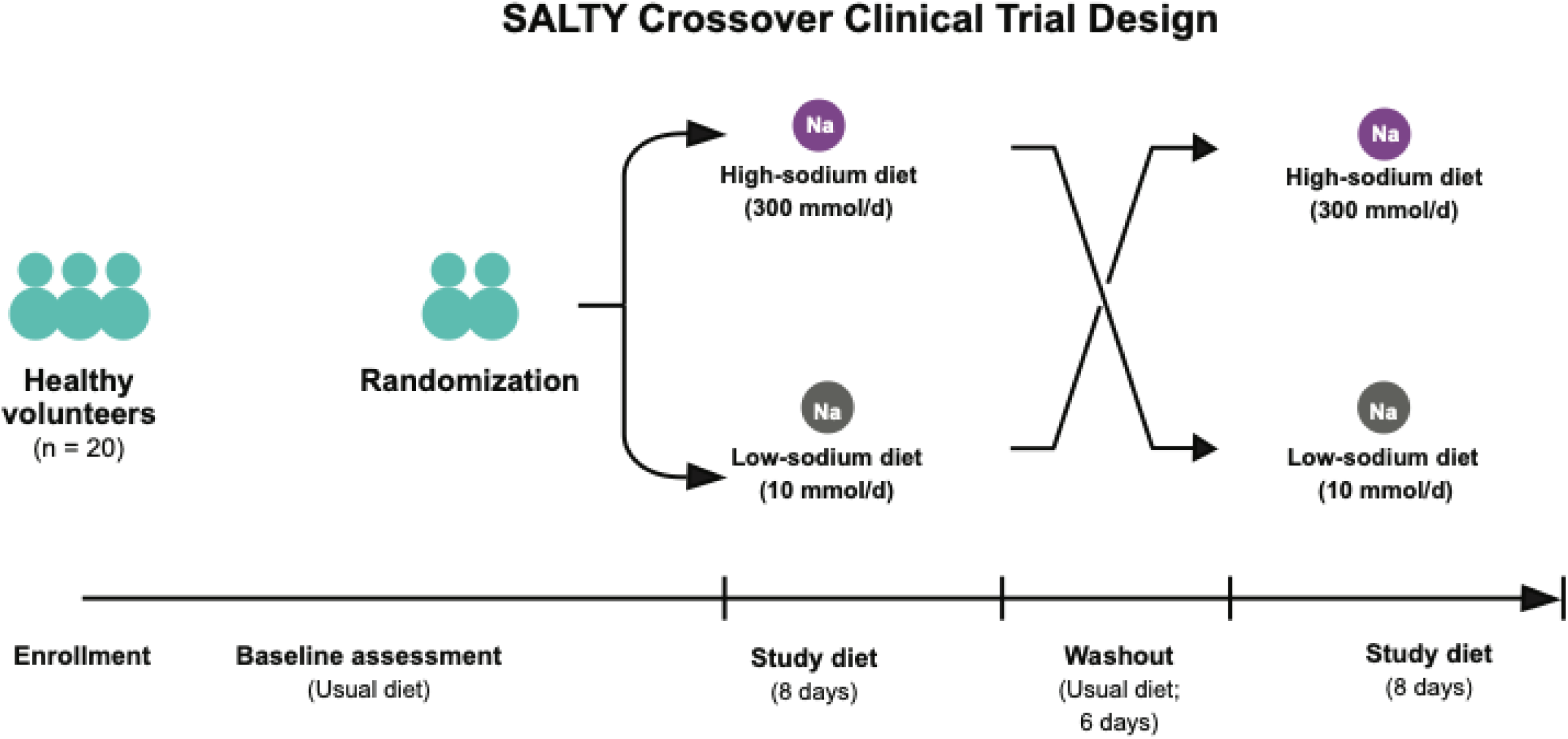
Study design of the SALTY crossover clinical trial. Schematic showing the randomized crossover design with participants consuming controlled low sodium (10 mmol/day) and high sodium (300 mmol/day) diets for 8 days each, separated by a 6-day washout period. All meals were provided by the research team to ensure dietary adherence. Standardized blood pressure measurements made in triplicate, 24-hour urine collection, and plasma sampling were performed at the end of each dietary period.

### Controlled Dietary Interventions

All meals were provided by the University of Michigan’s Metabolic Kitchen using standardized menus. Participants consumed low-sodium (10 mmol/day) and high-sodium (300 mmol/day) diets for 8 days each, separated by a 6-day washout period. Both diets contained the same foods during the high- and low-sodium diets, with additional table salt added to the foods during the high-sodium dietary phase. This approach ensured that macronutrient composition and potassium content were held constant. Participants were randomized to diet sequence (high-sodium then low-sodium, or low-sodium then high-sodium). Adherence with the diet was assessed by 24-hour urinary sodium measurement.

### Blood Pressure Measurements

Blood pressure was measured using the Omron HEM-907XL automated oscillometric device used in the SPRINT clinical trial.^16^ After a 5-minute seated rest period, three automated measurements were taken 1 minute apart using appropriate cuff sizes. The final two measurements were averaged for analysis. All measurements were taken at consistent times at the end of each dietary period in the fasting state.

### Proteomics Assays

SOMAmers (Slow Off-rate Modified Aptamers) are protein affinity reagents. Each SOMAmer reagent is highly specific to an epitope region of a respective cognate protein. The quantified aptamers therefore measure the relative concentration of a protein in a plasma sample. Plasma samples collected at the end of each dietary period were analyzed using SomaLogic’s SOMAscan 7K platform at SomaLogic Inc., Boulder, Colorado. The SOMAscan 7K platform measures 6,627 unique human proteins using 7,228 SOMAmers, modified single-stranded DNA aptamers with established analytical precision and reproducibility. Two independent aptamers targeting different SVEP1 domains provided internal validation.

### Statistical Analysis

#### Statistical Approach

The crossover design eliminates between-subject variability, increasing power to detect within-subject changes. Each participant provides paired observations under both dietary conditions, making this statistically more efficient than a parallel-group design with equivalent total observations. The effect of dietary sodium on clinical parameters such as weight was analyzed using linear mixed models accounting for treatment, period, and sequence effects with random subject intercepts. We used the Pearson correlation coefficient for protein-blood pressure relationships. Salt sensitivity was classified based on mean arterial pressure (MAP) changes: inverse salt sensitivity (≥5 mmHg decrease), salt resistance (<5 mmHg change), and salt sensitivity (≥5 mmHg increase).

#### Proteomic Analysis

For 19 of the 20 participants, plasma was available for proteomics. Plasma protein levels from each dietary period were analyzed using log2 transformation to normalize distributions and enable fold-change calculations. For each protein, we analyzed change in plasma protein level using linear mixed effect models to test for differences between high- and low-sodium dietary periods after adjusting for sequence (high-then-low versus low-then-high). We used the ‘lmerTest’ package in R to construct these models in the following fashion: log2_value ∼ treatment + period + sequence + (1|subject). This approach models the effect of dietary sodium (high versus low) on each protein, accounting for potential period and sequence effects in the crossover design.

The SOMAscan 7K platform measured 7,288 aptamers targeting 6,627 unique human proteins. When multiple aptamers targeted the same protein, each was analyzed independently to provide technical validation. All aptamer measurements were ranked by p-value from the linear mixed effect model. Benjamini-Hochberg false discovery rate (FDR) correction was applied for multiple comparison correction in proteomic analyses. Proteins with FDR-adjusted q-values <0.05 were considered significant after multiple comparison correction. Secondary analyses examined correlations between protein changes and blood pressure responses using Pearson correlation coefficients. All analyses were performed using R version 4.5.1 with SomaDataIO (version 6.3.0) for processing SOMAscan data.

#### Reactome Pathway Analysis

To identify biological pathways significantly enriched among proteins showing differential expression between high- and low-sodium diets, we performed pathway enrichment analysis using the Reactome database implemented through the clusterProfiler package in R. Gene symbols of significantly differentially expressed proteins (FDR < 0.05) were analyzed against a reference background of all detected and annotated proteins in the dataset. The enrichPathway function automatically filtered the background to include only genes present in Reactome pathways (n = 4,403 genes from 6,346 unique annotated genes). Statistical significance was assessed using the hypergeometric test with Benjamini-Hochberg correction for multiple testing, and pathways with adjusted p-values (q-values) < 0.05 were considered significantly enriched.

For each pathway, we calculated several enrichment metrics: gene ratio (proportion of input genes mapping to the pathway), background ratio (proportion of all background genes in the pathway), rich factor (gene ratio divided by background ratio as a measure of enrichment strength), fold enrichment (ratio of observed to expected gene overlap), Z-score (standardized enrichment score accounting for pathway size), p-value (hypergeometric test), and q-value (Benjamini-Hochberg adjusted p-value).

## Results

### Physiological Responses to Controlled Sodium Loading

Twenty participants (**Table 1**) completed the SALTY study, with adequate dietary adherence confirmed by 24-hour urine sodium monitoring (**Figure 2**). The sodium loading intervention successfully engaged established sodium regulatory pathways, with plasma renin activity being 9.8 ng/mL/h during high sodium intake versus 36.5 ng/mL/h during low sodium intake (p<0.001), and serum aldosterone being 9.4 ng/dL during high sodium intake versus 37.1 ng/dL during low sodium intake (p<0.001) (**Table 2**). These expected hormonal responses confirmed effective sodium loading and provided context for interpreting novel proteomic findings.

**Figure 2.**
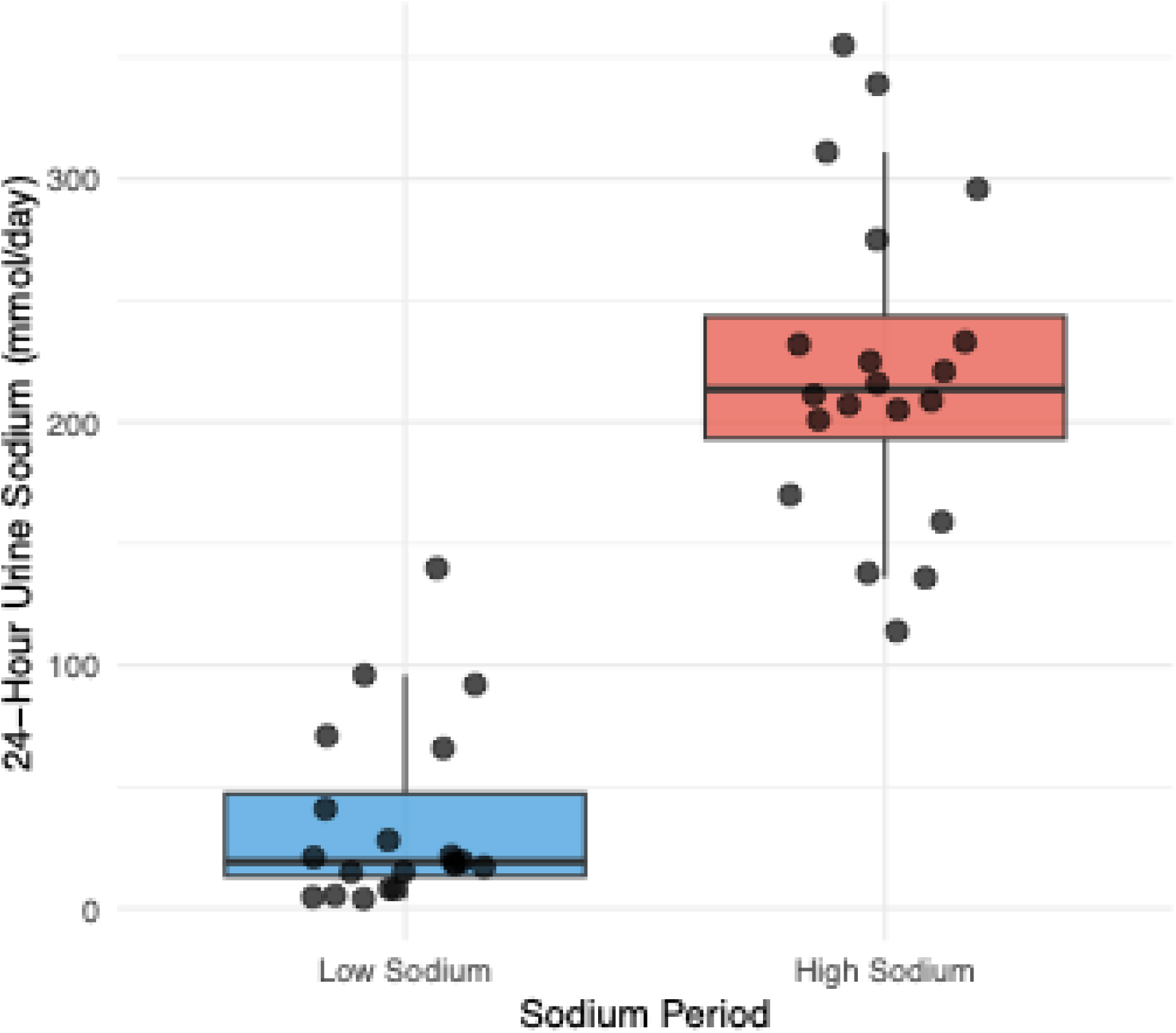
24-Hour urinary sodium excretion by dietary period. Box plots showing individual participant’s 24-hour urinary sodium excretion during low and high sodium diets. Each dot represents one participant during one dietary period. For samples with sodium concentration below the detection limit of 10 mmol/L, the 24-hour sodium excretion was calculated using an imputed concentration of 5 mmol/L (LLOD/2) multiplied by the participant’s 24-hour urine volume (n=7, all during low-sodium diet). Analysis included all N=20 participants who completed the SALTY study. P < 0.001 by linear mixed model accounting for treatment, period, sequence, and random subject effects.

**Table 1.**
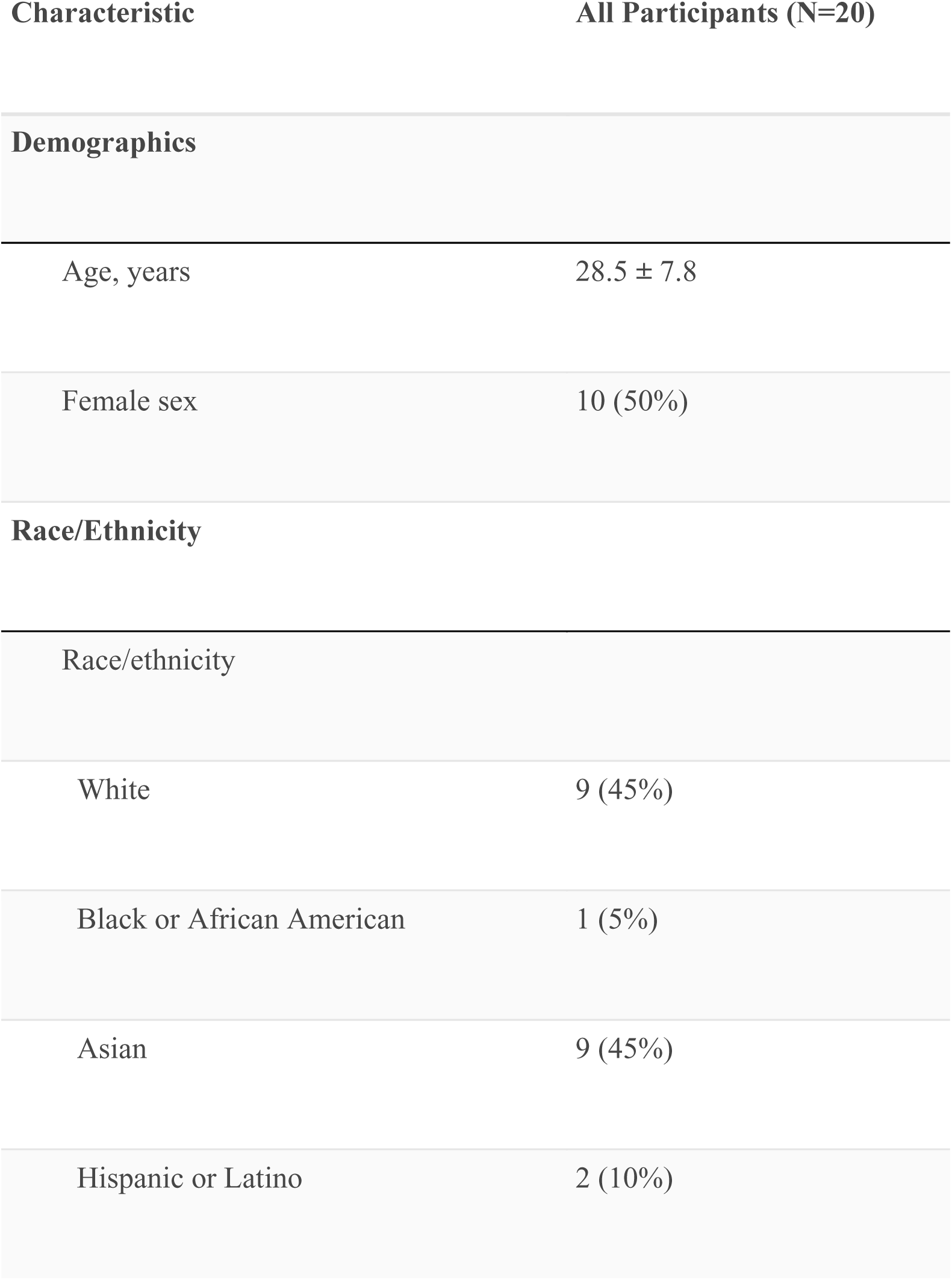
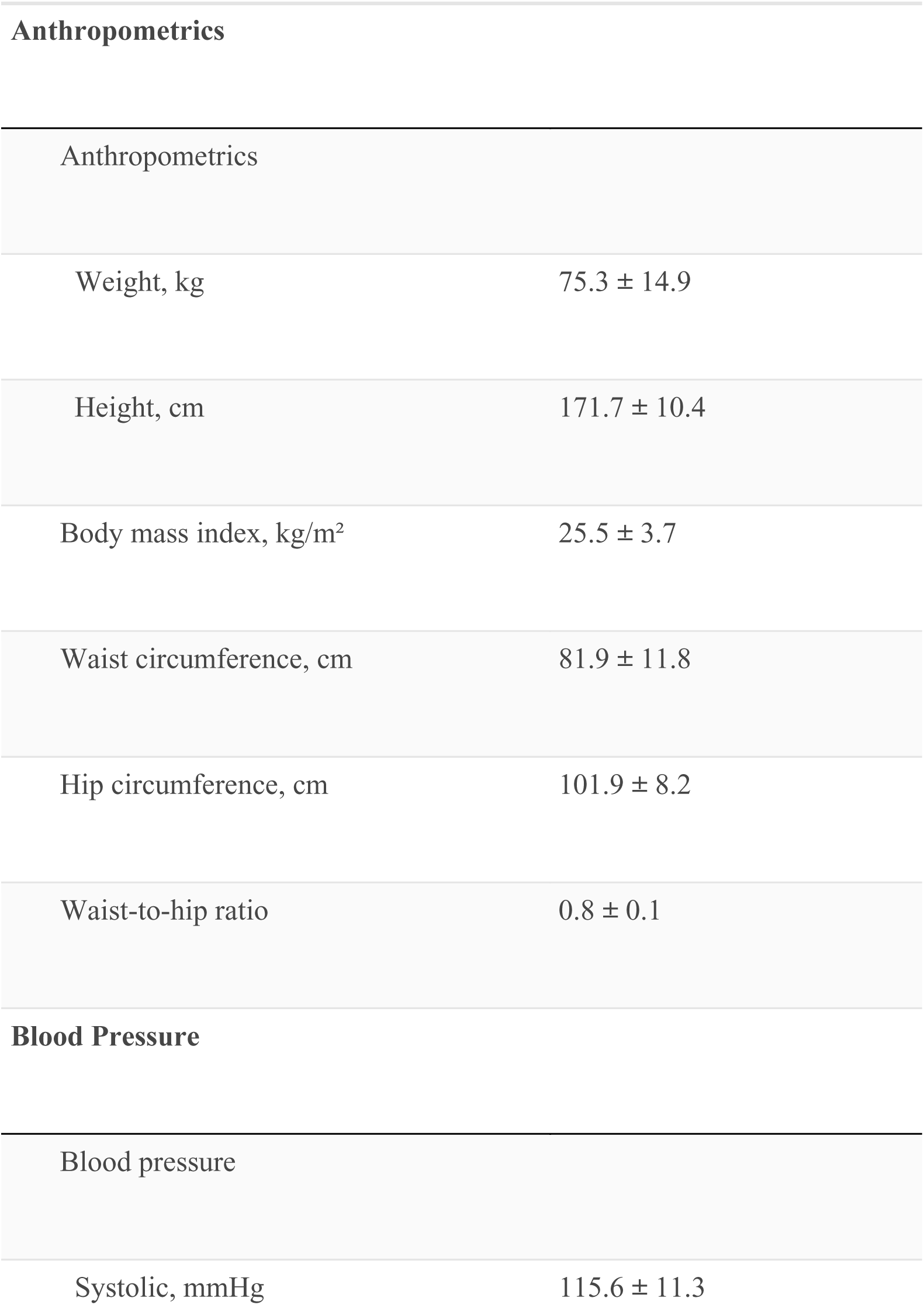
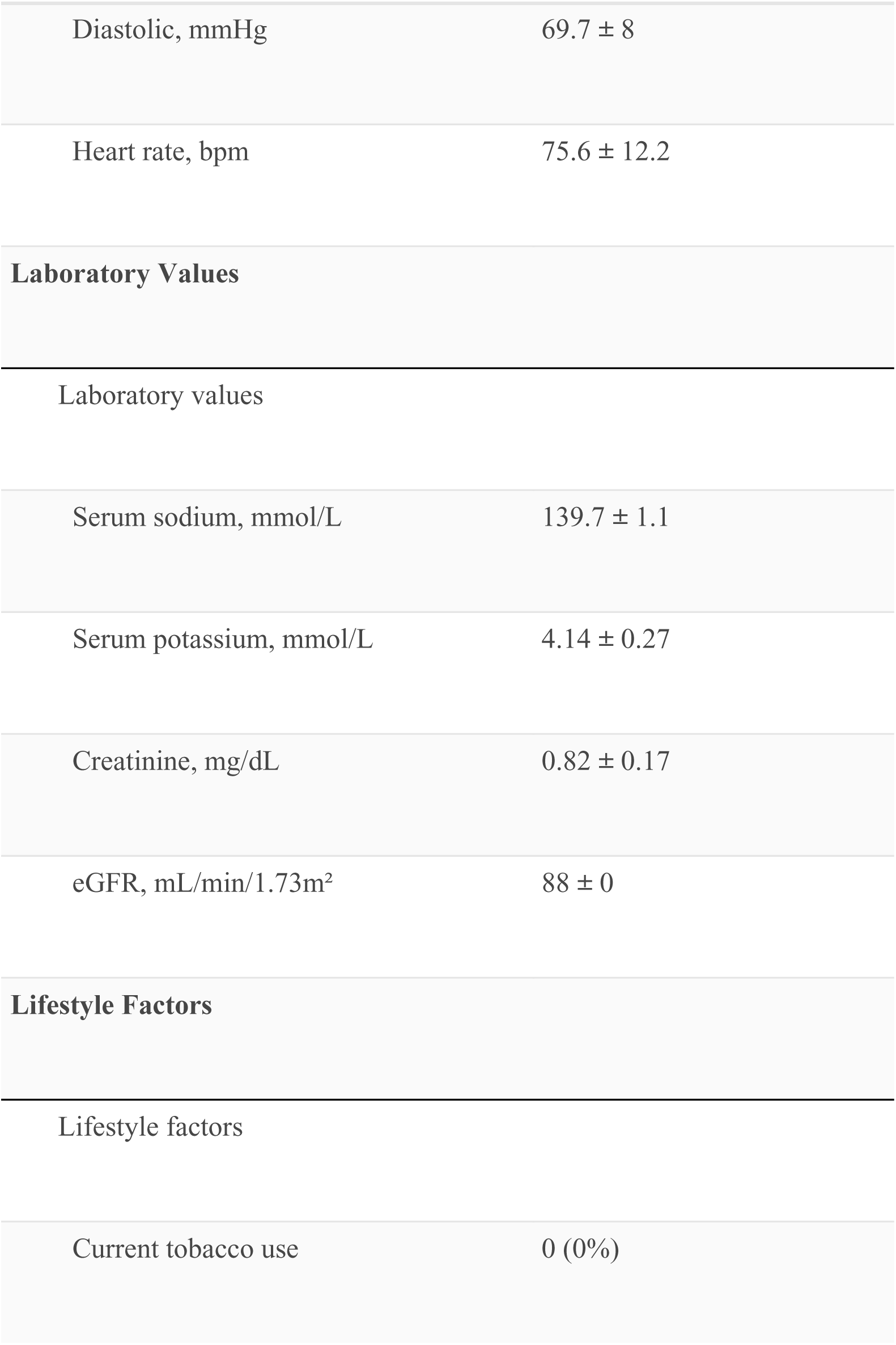
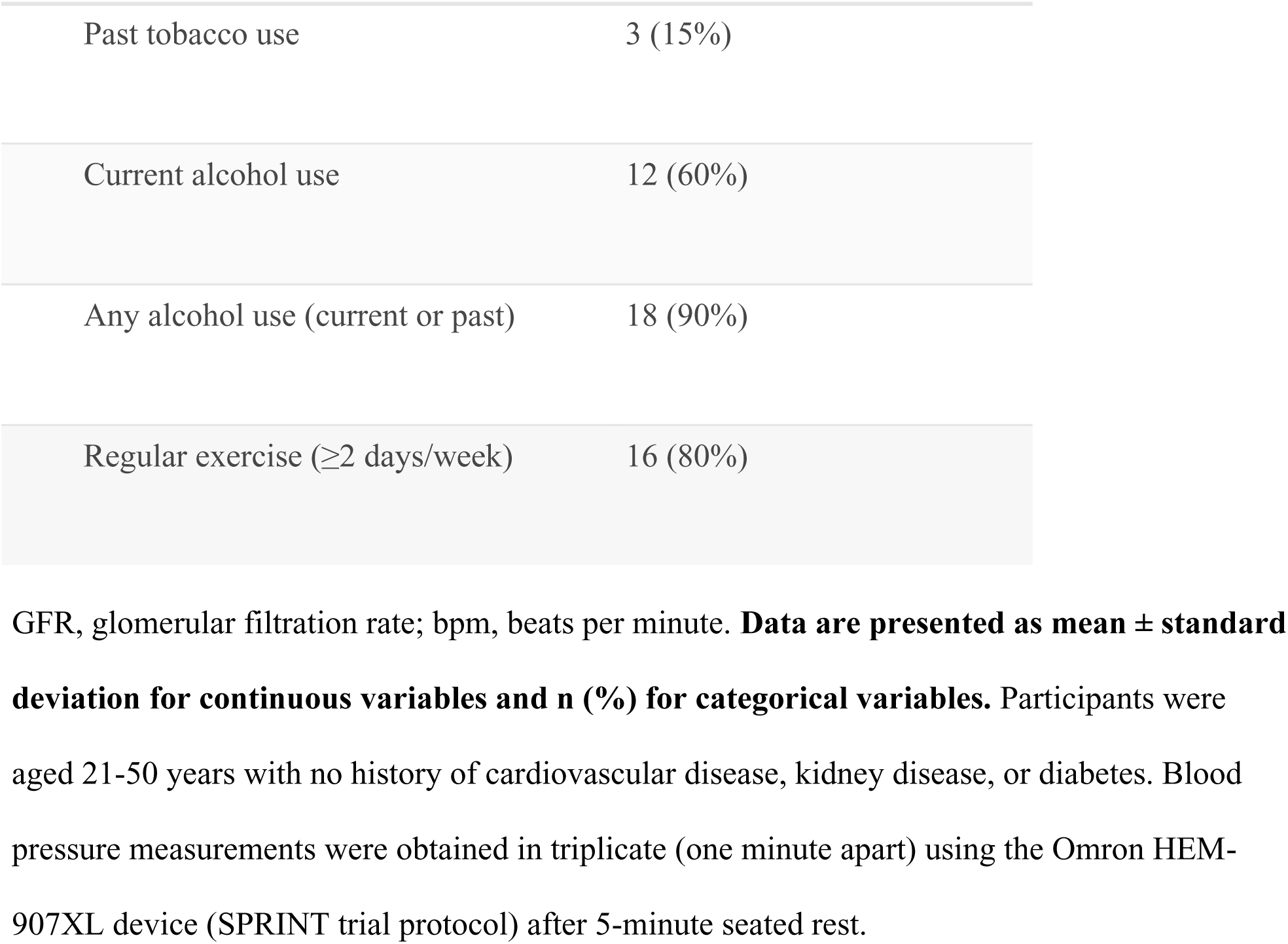
SALTY clinical trial baseline participant characteristics (N=20).

| Characteristic | All Participants (N=20) |
| --- | --- |
| Demographics |  |
| Age, years | 28.5 ± 7.8 |
| Female sex | 10 (50%) |
| Race/Ethnicity |  |
| Race/ethnicity |  |
| White | 9 (45%) |
| Black or African American | 1 (5%) |
| Asian | 9 (45%) |
| Hispanic or Latino | 2 (10%) |
| Anthropometrics |  |
| Weight, kg | 75.3 ± 14.9 |
| Height, cm | 171.7 ± 10.4 |
| Body mass index, kg/m² | 25.5 ± 3.7 |
| Waist circumference, cm | 81.9 ± 11.8 |
| Hip circumference, cm | 101.9 ± 8.2 |
| Waist-to-hip ratio | 0.8 ± 0.1 |
| Blood Pressure |  |
| Blood pressure |  |
| Systolic, mmHg | 115.6 ± 11.3 |
| Diastolic, mmHg | 69.7 ± 8 |
| Heart rate, bpm | 75.6 ± 12.2 |
| Laboratory Values |  |
| Laboratory values |  |
| Serum sodium, mmol/L | 139.7 ± 1.1 |
| Serum potassium, mmol/L | 4.14 ± 0.27 |
| Creatinine, mg/dL | 0.82 ± 0.17 |
| eGFR, mL/min/1.73m<sup>2</sup> | 88 ± 0 |
| Lifestyle Factors |  |
| Lifestyle factors |  |
| Current tobacco use | 0 (0%) |
| Past tobacco use | 3 (15%) |
| Current alcohol use | 12 (60%) |
| Any alcohol use (current or past) | 18 (90%) |
| Regular exercise ($\geq 2$ days/week) | 16 (80%) |

GFR, glomerular filtration rate; bpm, beats per minute. **Data are presented as mean ± standard** **deviation for continuous variables and n (%) for categorical variables.** Participants were aged 21-50 years with no history of cardiovascular disease, kidney disease, or diabetes. Blood pressure measurements were obtained in triplicate (one minute apart) using the Omron HEM- 907XL device (SPRINT trial protocol) after 5-minute seated rest.

**Table 2.** Effect of dietary sodium intake on clinical and laboratory parameters.

| Parameter | Units | Low | High | Mean Difference (95% CI) | P value | n |
| --- | --- | --- | --- | --- | --- | --- |
|  |  | Sodium | Sodium |  |  |  |
| Blood Pressure Responses |  |  |  |  |  |  |
| Systolic blood pressure | mmHg | 115 ± 10 | 112 ± 10 | -2.5 (-5.2 to 0.1) | 0.060 | 20 |
| Diastolic blood pressure | mmHg | 70 ± 8 | 67 ± 8 | -2.6 (-4.7 to -0.6) | 0.014 | 20 |
| Mean arterial pressure | mmHg | 84.8 ± 8.3 | 82.1 ± 7.6 | -2.62 (-4.58 to -0.65) | 0.012 | 20 |
| Markers of Change in Plasma Volume |  |  |  |  |  |  |

| Parameter | Units | Low<br>Sodium | High<br>Sodium | Mean Difference (95% CI) | P value | n |
| --- | --- | --- | --- | --- | --- | --- |
| Body weight | kg | 74.3 ± 14.0 | 75.7 ± 14.3 | 1.36 (0.95 to 1.77) | <0.001 | 20 |
| Sodium Intervention Validation |  |  |  |  |  |  |
| 24-hour urine sodium | mmol/day | 52 ± 41 | 223 ± 66 | 174 (134 to 214) | <0.001 | 20 |
| Serum sodium | mmol/L | 138.8 ± 1.1 | 139.9 ± 1.4 | 1.05 (0.53 to 1.57) | <0.001 | 20 |
| Potassium |  |  |  |  |  |  |
| 24-hour urine<br>potassium | mmol/day | 50 ± 22 | 60 ± 25 | 9 (-4 to 22) | 0.155 | 20 |
| Serum potassium | mmol/L | 4.3 ± 0.2 | 4.2 ± 0.3 | -0.10 (-0.26 to 0.05) | 0.175 | 20 |
| **Hormones** |  |  |  |  |  |  |
| Plasma renin activity | ng/mL/h | 36.5 ± 27.8 | 9.8 ± 5.8 | -26.8 (-37.8 to -15.8) | <0.001 | 20 |
| Serum aldosterone | ng/dL | 37.1 ± 24.1 | 9.4 ± 4.9 | -27.7 (-38.4 to -17.0) | <0.001 | 20 |

Data are presented as mean ± SD. P values are from linear mixed models accounting for treatment, period, and sequence effects with random subject intercepts.

**Table 3.**
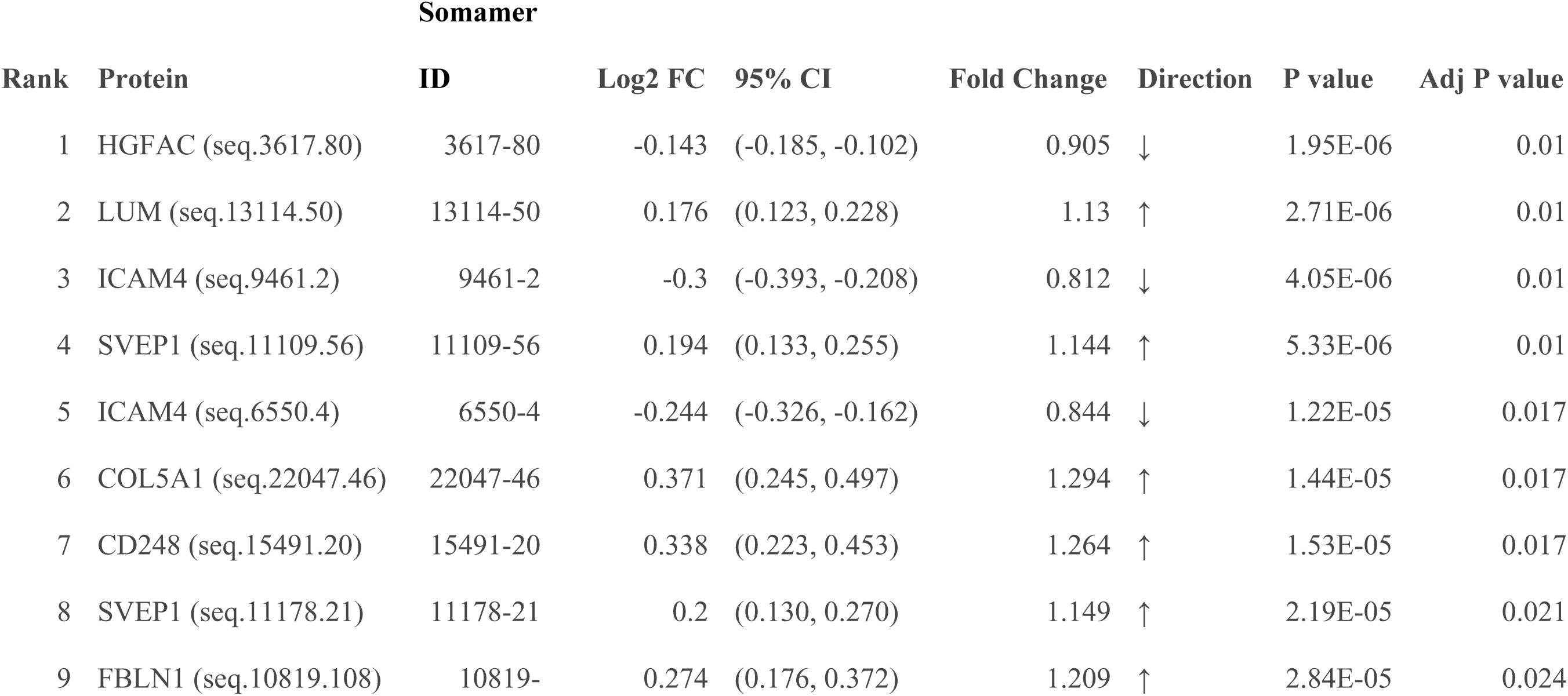
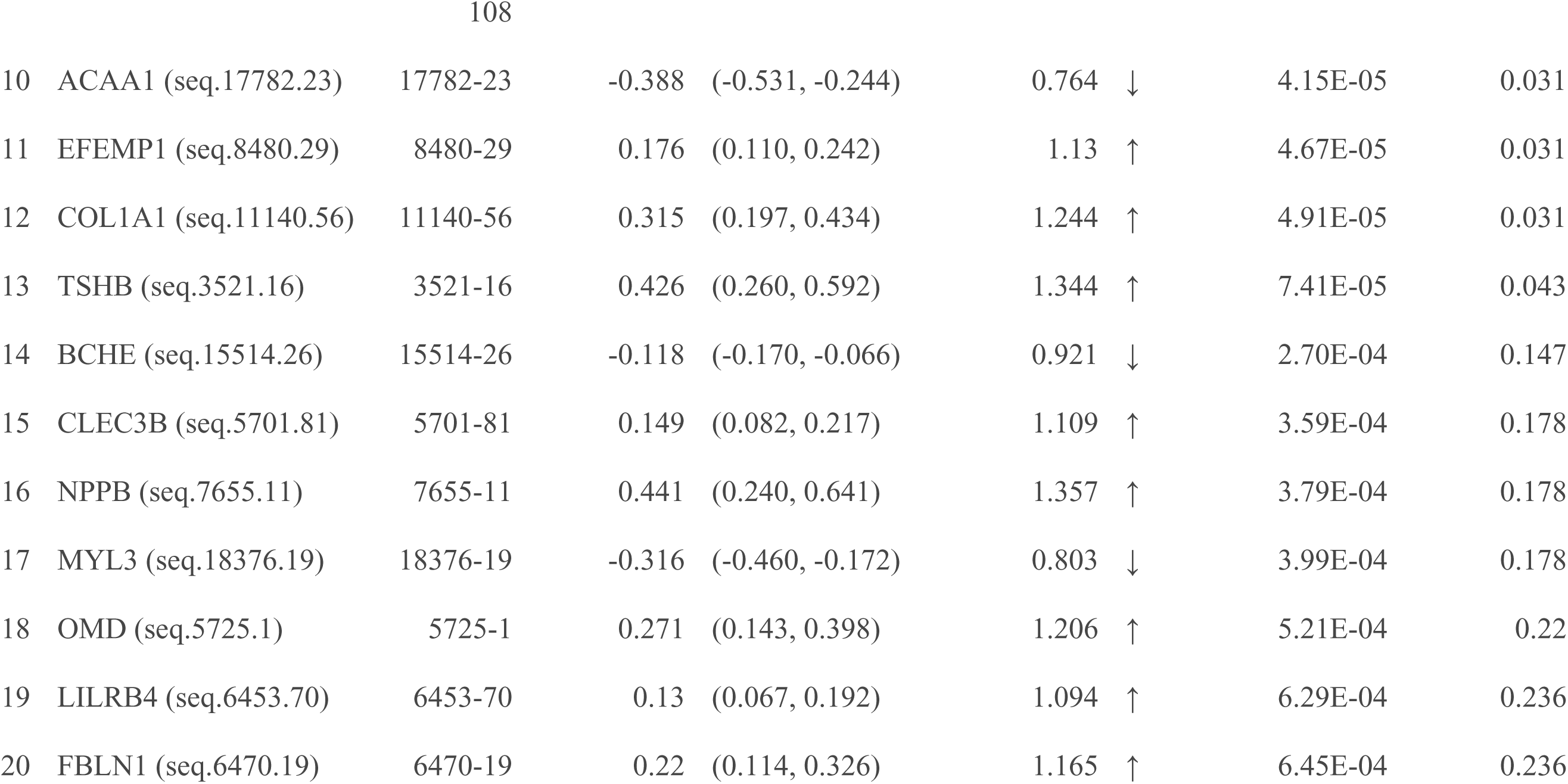
Top 20 plasma proteins responding to dietary sodium intake (n=19). Proteins ranked by statistical significance from linear mixed models testing the effect of high (6,900 mg/day) versus low (230 mg/day) sodium diet. Log2 fold change and 95% confidence intervals represent the treatment effect adjusted for period and sequence effects with random subject intercepts. P-values are unadjusted; adjusted p-values use Benjamini-Hochberg correction for multiple testing across ∼7,000 proteins. Direction arrows (↑/↓) indicate increased or decreased expression with high sodium intake.

Participants demonstrated significant weight difference during high-sodium versus low-sodium intake, averaging +1.4 kg (95% CI: 0.93-1.79 kg, p=8.85×10^-7^) during high-sodium diet, consistent with volume expansion during sodium loading. Despite the observed weight gain consistent with volume expansion, blood pressure responses varied markedly among participants. Both diastolic blood pressure and MAP were significantly lower during high sodium intake (diastolic blood pressure: 67.0±7.5 vs 69.7±8.0 mm Hg, p=0.014; MAP: 82.2±7.6 versus 84.8±8.3 mm Hg, p=0.029). When participants were classified by MAP response (using a ≥5 mmHg threshold), only 5% (1/20) showed traditional salt sensitivity, while 35% (7/20) exhibited inverse salt sensitivity—blood pressure decrease during high sodium diet—and the remaining 60% (12/20) were salt resistant (**Figure 3**).

**Figure 3.**
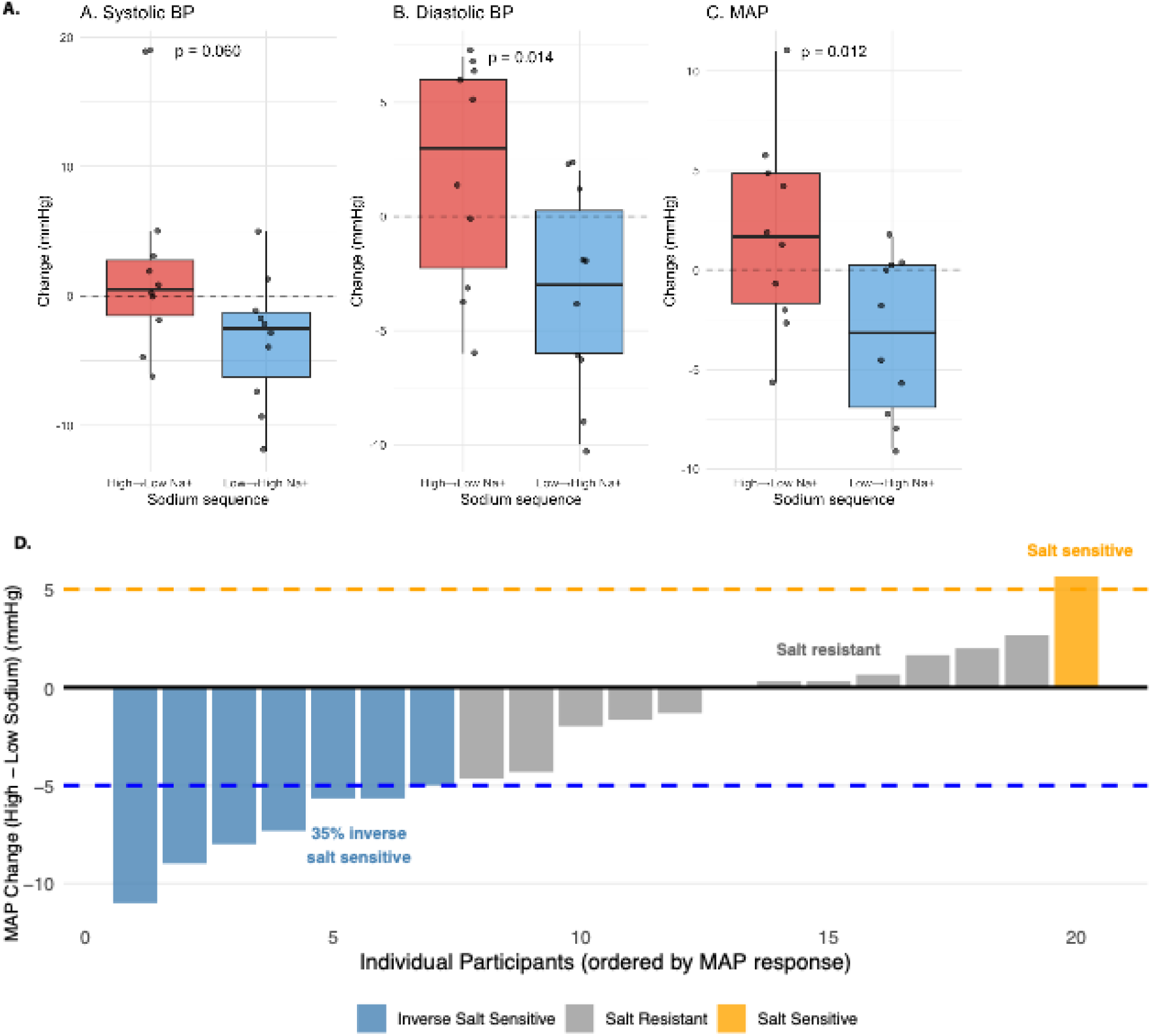
Individual blood pressure responses to sodium loading. (**A-C**) Box plots showing changes in systolic blood pressure (A), diastolic blood pressure (B), and mean arterial pressure (C) between high and low sodium dietary periods, grouped by randomization sequence. Each dot represents one participant’s response. Horizontal dashed lines indicate no change. P-values shown are from linear mixed models accounting for treatment, period, sequence, and random subject effects. (**D**) Waterfall plot showing individual mean arterial pressure responses to high versus low sodium diet, ranked from most inverse salt sensitive to most salt sensitive. Salt sensitivity phenotypes were defined as: inverse salt sensitive (≥5 mmHg MAP decrease with high sodium), salt resistant (<5 mmHg change), and salt sensitive (≥5 mmHg MAP increase with high sodium). Analysis included all SALTY study completers (N=20). Distribution: 35% inverse salt sensitive, 60% salt resistant, 5% salt sensitive.

### SVEP1 Emerges as a Prominent Proteomic Response

Among all 7,288 aptamers against proteins analyzed, SVEP1 (Sushi, von Willebrand factor type A, EGF and pentraxin domain containing 1) demonstrated the 4^rd^ most significant response to sodium loading (**Figure 4**).

**Figure 4.**
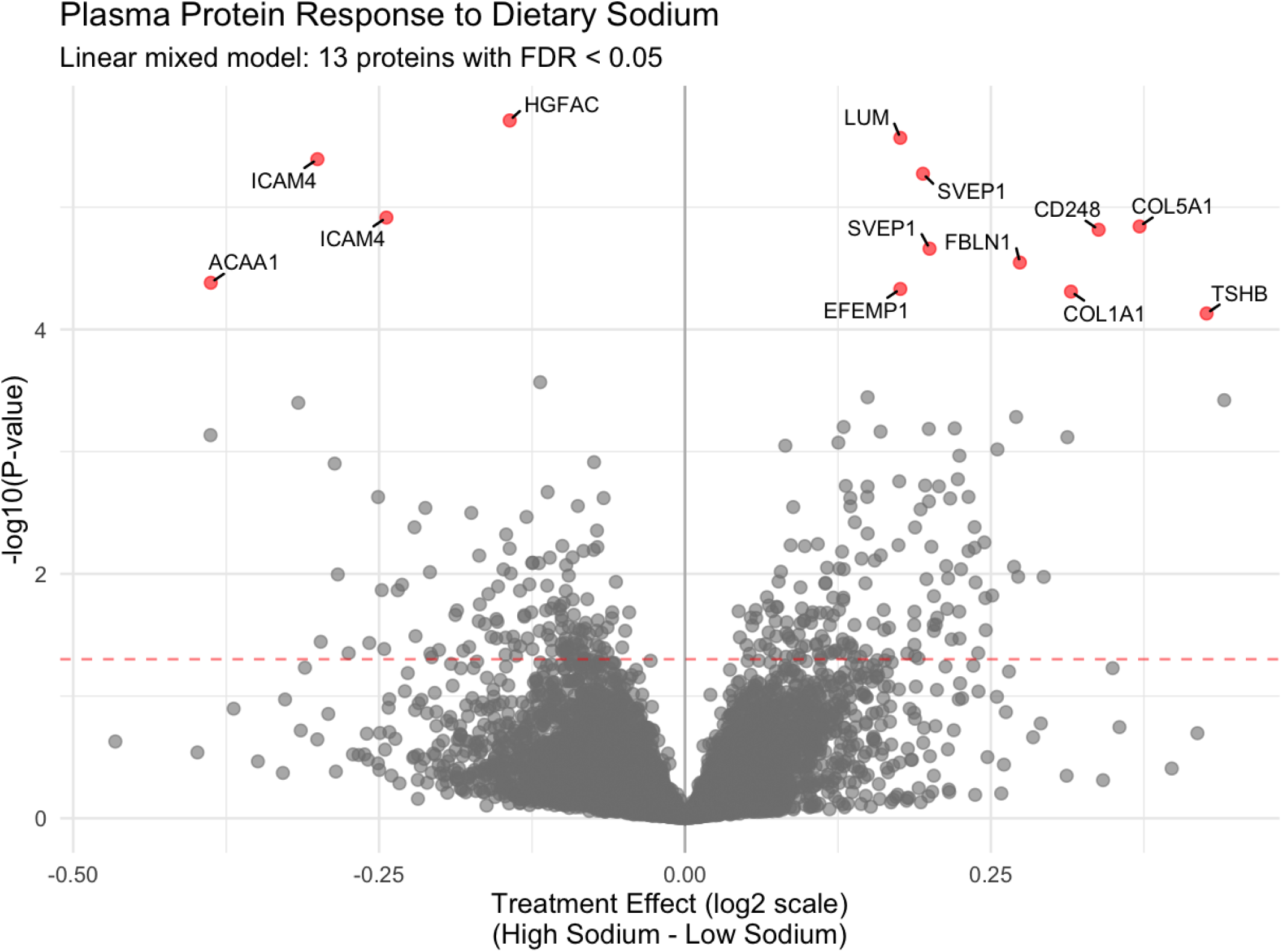
Plasma protein response to dietary sodium loading. Volcano plot showing the relationship between effect size (x-axis) and statistical significance (y-axis) for plasma protein changes in response to low and high sodium diets. Each point represents one protein. When multiple aptamers targeted the same protein, the aptamer with the lowest p-value was selected to represent that protein. Red points indicate proteins with significant changes after Benjamini-Hochberg correction for multiple testing (FDR q < 0.05). The x-axis shows the log2 fold-change in protein levels comparing high sodium to low sodium dietary periods. The y-axis shows the negative log10 of the p-value, with higher values indicating greater statistical significance. The horizontal dashed line indicates the nominal significance threshold (p = 0.05). Six proteins showed significant changes: COL5A1, CD248, LUM, and SVEP1 increased with high sodium intake, while ICAM4 and HGFA decreased with high sodium intake. Analysis included n = 19 SALTY participants who for whom plasma was available.

Two independent aptamers targeting different SVEP1 domains ranked 4th (p=5.32×10^-6^) and 8th (p=2.19×10^-5^) in statistical significance out of all the aptamers analyzed (**Figure 5**), with high correlation between measurements by these two aptamers directed at SVEP1 (R=0.93, p<0.001). The effect sizes for the two SVEP1-directed aptamers were almost identical, although they detect distinct domains of the protein. After FDR correction of the p-values for multiple comparisons, both aptamers directed at SVEP1 domains remained significant (FDR q=0.01 and 0.02, respectively).

**Figure 5.**
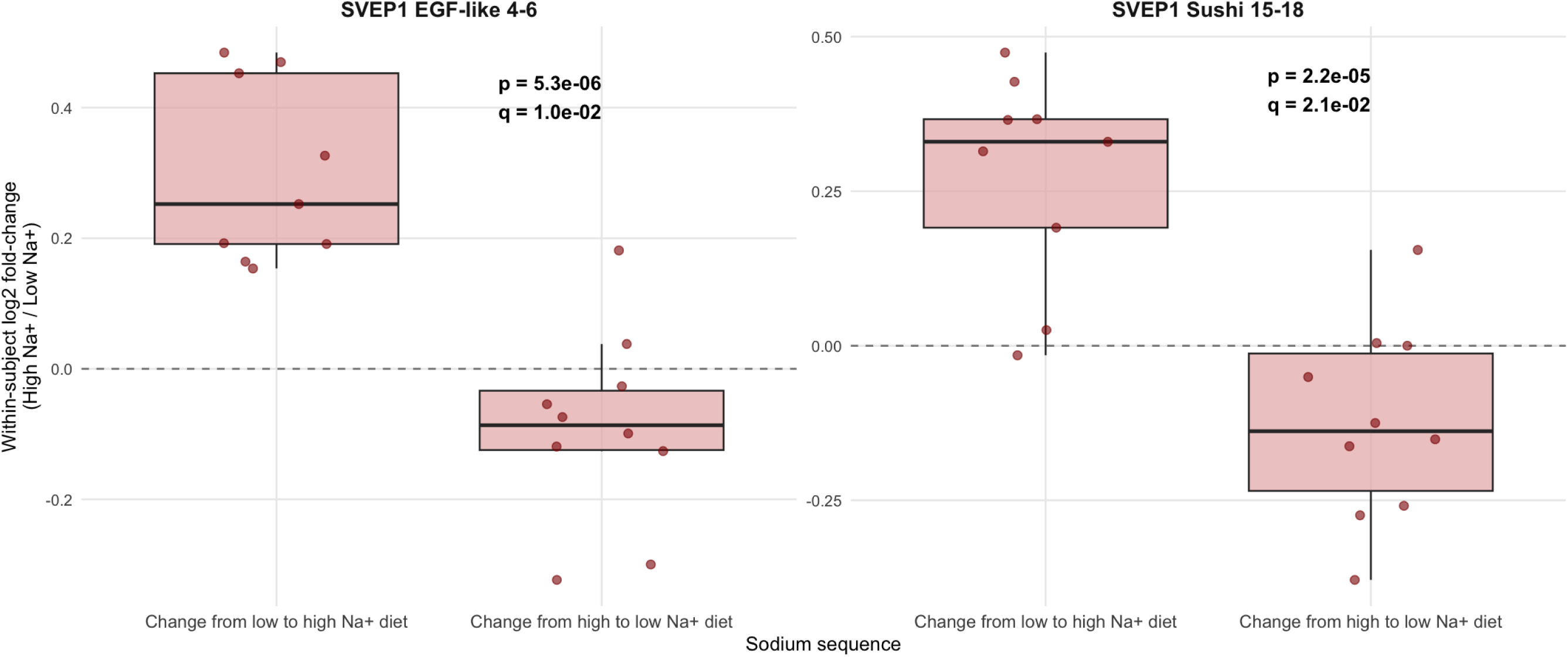
SVEP1 protein responses (both aptamers) comparing high versus low sodium diet. Box plots showing within-subject log2 fold-changes in SVEP1 protein levels (high sodium / low sodium) for two independent aptamers targeting different functional domains of SVEP1: Sushi domains 15-18 (aptamer 11109-56) and EGF-like domains 4-6 (aptamer 11178-21). Data are shown by sequence group accounting for the crossover design. Each dot represents one participant’s response. Horizontal dashed lines indicate no change (log2 fold-change = 0). Both aptamers demonstrated highly significant increases in SVEP1 levels during high sodium intake (p = 1.7e-05 and p = 6.1e-05, respectively), with consistent upregulation across different functional domains confirming the protein-level finding from the discovery analysis (protein-level FDR q = 0.033). Analysis included N = 19 participants who completed both the SALTY dietary crossover study and for whom plasma was available for proteomics.

### Change in SVEP1 Correlates Inversely with Blood Pressure Response to Dietary Sodium Loading

SVEP1 upregulation correlated inversely with MAP changes during sodium loading (**Figure 6**, r=-0.50, p=0.028), indicating that greater SVEP1 responses were associated with greater blood pressure reductions or smaller increases. Among all aptamers analyzed, SVEP1’s correlation with blood pressure changes ranked 52nd (top 0.7%) in statistical significance, substantially outranking canonical markers of sodium metabolism such as renin, BNP, and NT-ProBNP.

**Figure 6.**
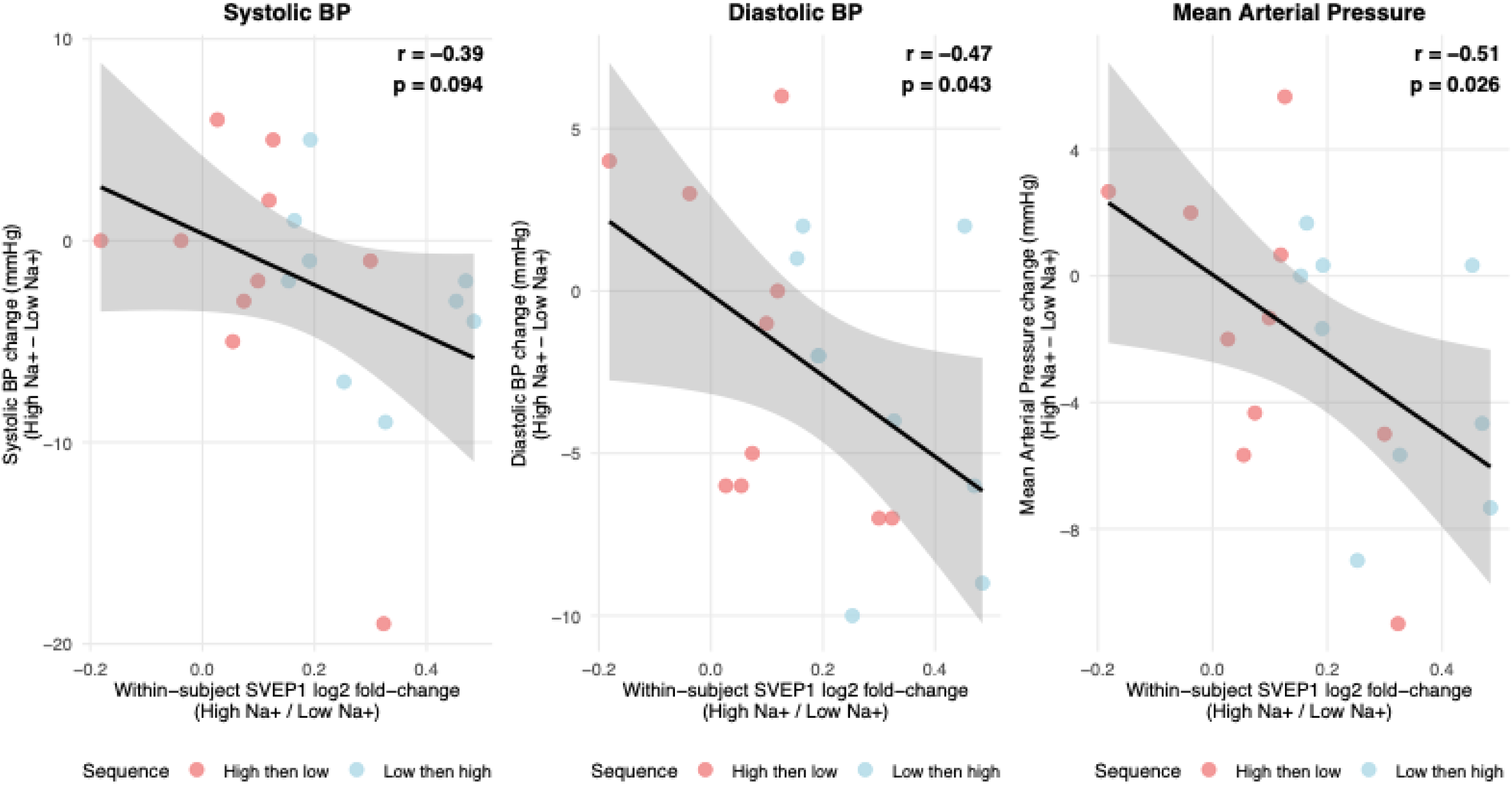
Correlation between SVEP1 changes and blood pressure changes between low versus high sodium diet. Scatter plots showing the relationship between within-subject SVEP1 log2 fold-changes (Sushi domains 15-18) and blood pressure changes for systolic blood pressure (left), diastolic blood pressure (center), and mean arterial pressure (right). Each point represents one participant’s paired response to the dietary sodium intervention. Colors indicate crossover sequence: coral for participants who received high sodium first, blue for those who received low sodium first. Black lines show linear regression with 95% confidence intervals. A larger increase in SVEP1 levels with high sodium intake were associated with smaller blood pressure increases or larger blood pressure decreases, consistent with a protective cardiovascular effect. The inverse correlations were statistically significant for diastolic blood pressure (r = -0.47, p = 0.043) and mean arterial pressure (r = -0.51, p = 0.026), with systolic blood pressure showing a similar trend (r = -0.39, p = 0.094). Analysis included the n=19 SALTY crossover study participants for whom plasma proteomics were available.

Individuals exhibiting inverse salt sensitivity demonstrated approximately 2-fold higher SVEP1 increases compared to salt-resistant participants, consistent with SVEP1 serving as a biomarker for identifying individuals with enhanced adaptive capacity to sodium challenges.

### Change in NT-ProBNP Correlates Strongly with Change in SVEP1 in the Transition Between High- and Low-sodium Diets

Plasma NT-ProBNP levels correlated with SVEP1 levels (r = 0.69, p < 0.001). We further investigated the possibility that changes in the plasma volume regulate SVEP1. We analyzed the correlation between the change in NT-ProBNP and change in SVEP1 during the transition between the high- and low-sodium diets. The change in NT-ProBNP between the high- and the low-sodium dietary conditions correlated strongly with the change in SVEP1 (Pearson’s R=0.80, p<0.001; **Figure 7**).

**Figure 7.**
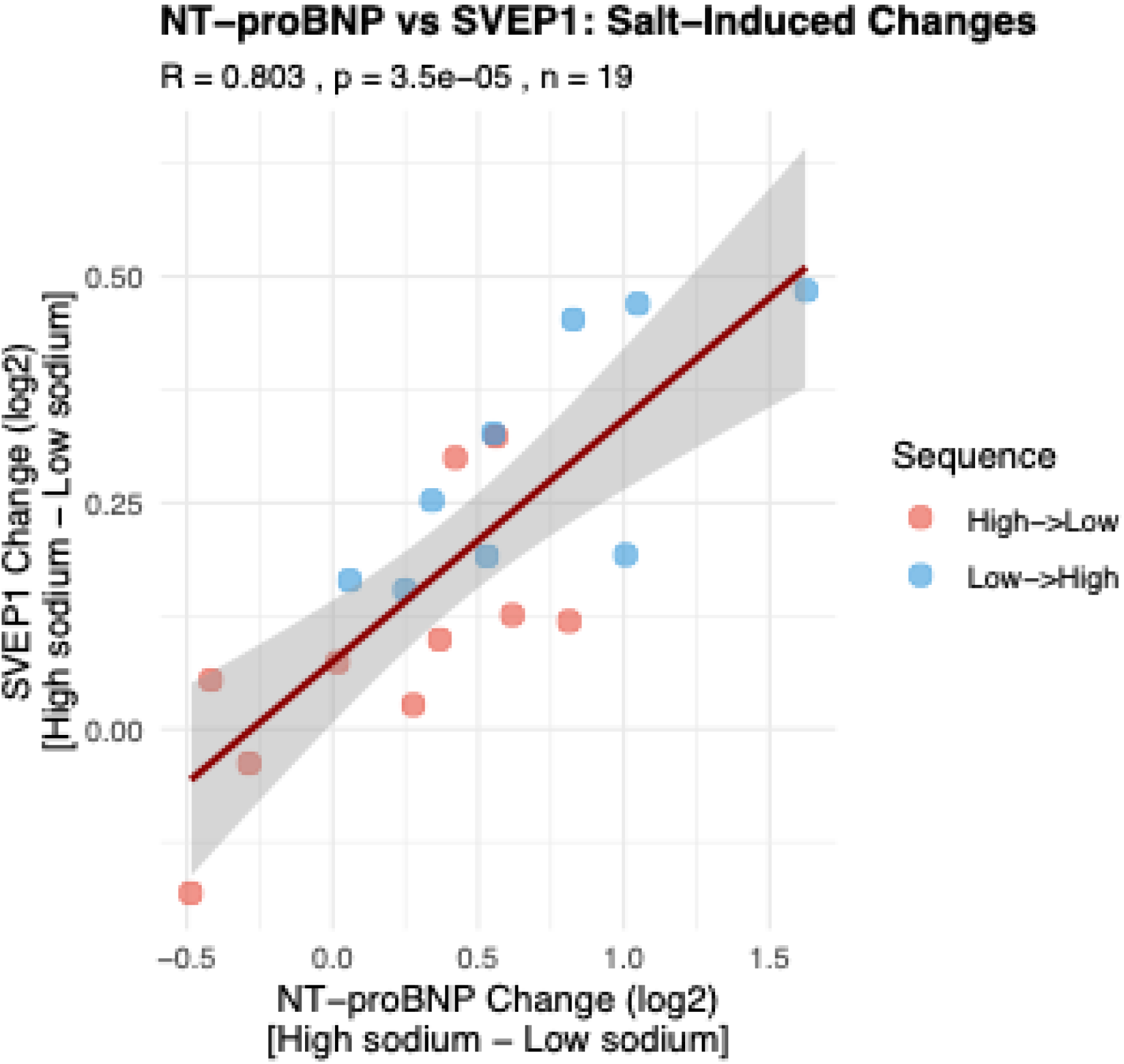
Relationship Between Sodium-induced Changes in Plasma NT-ProBNP and SVEP1. Correlation coefficient is per the Pearson method. The color of the dots indicates whether the first diet consumed by that participant was the high-sodium diet (red dots), or the low-sodium diet (blue dots).

### Coordinated Extracellular Matrix Response

Pathway enrichment analysis using the Reactome database identified 35 significantly enriched pathways (q < 0.05) among the 13 proteins showing differential expression between high and low salt treatments. The most significantly enriched pathway was ‘Extracellular matrix organization’ (6 proteins; fold enrichment = 14.6; q = 2.06 × 10⁻⁵), followed by ‘Integrin cell surface interactions’ (4 proteins; fold enrichment = 33.2; q = 5.69 × 10⁻⁵) and ‘ECM proteoglycans’ (3 proteins; fold enrichment = 28.8; q = 1.01 × 10⁻³). The analysis revealed a predominant enrichment of pathways related to extracellular matrix metabolism, with 8 of the top 10 pathways involving collagen processing, ECM organization, or cell-matrix interactions (**Figure 8**).

**Figure 8.**
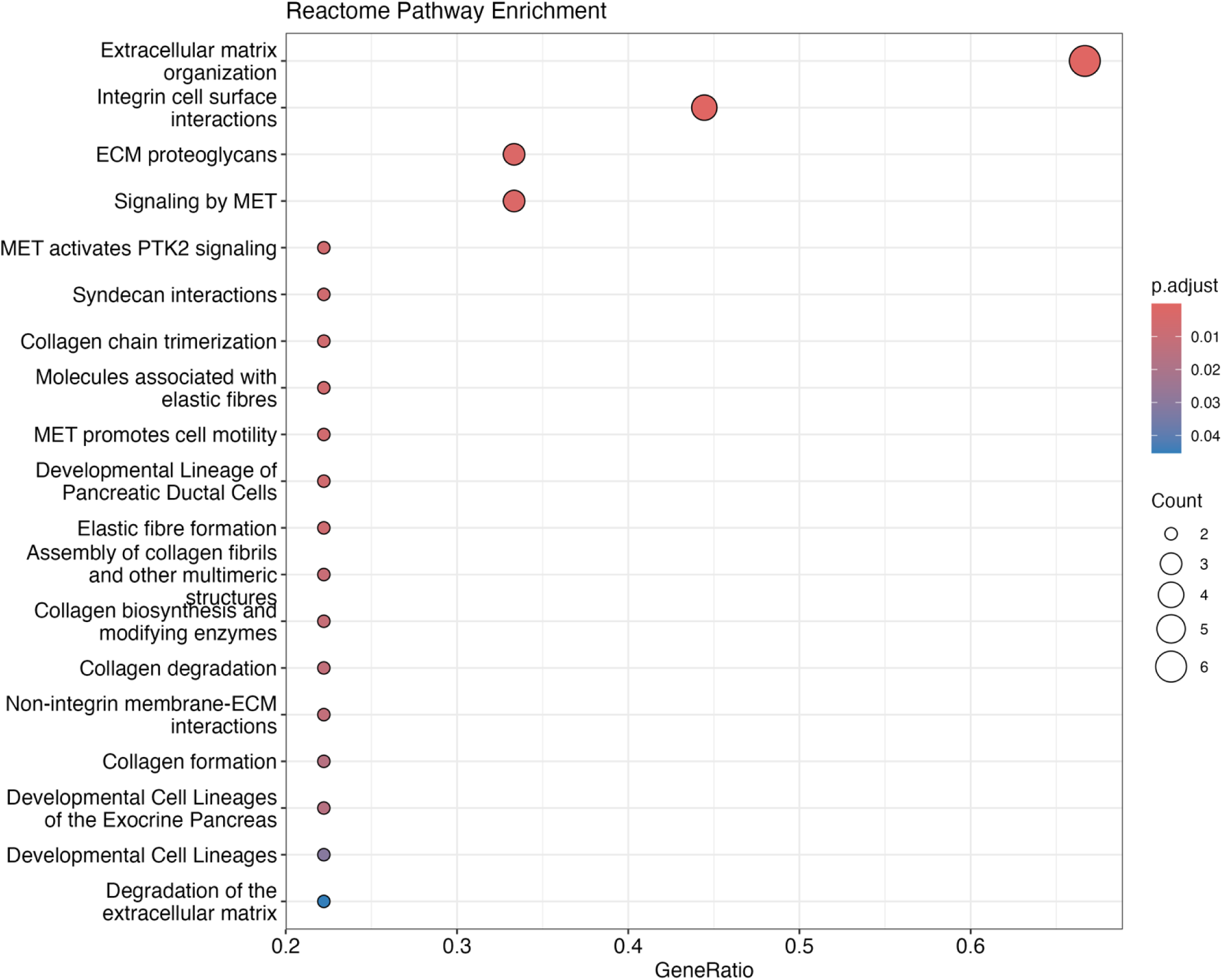
Reactome Pathway Enrichment Dot Plot for Differential Plasma Proteins. Dot plot showing the top Reactome pathways significantly enriched among proteins differentially expressed between high- and low-sodium intake arms. Each dot represents a Reactome pathway; the x-axis indicates gene ratio (proportion of significant proteins in the pathway), and the y-axis lists pathway names. Dot color represents the adjusted p-value for enrichment (Benjamini-Hochberg correction), and dot size reflects the number of significant proteins within each pathway. Pathways are ranked by statistical significance. This analysis provides a systems-level overview, contextualizing proteomic changes in biological processes and pathways altered by the dietary sodium intervention. Abbreviations: FDR, false discovery rate.

The enriched pathways clustered into several functional themes. Extracellular matrix-related processes were most prominent, including collagen metabolism pathways such as ‘Collagen chain trimerization’ (2 proteins; fold enrichment = 48.9; q = 3.23 × 10⁻³), ‘Collagen biosynthesis and modifying enzymes’ (2 proteins; fold enrichment = 28.0; q = 5.36 × 10⁻³), and ‘Collagen formation’ (2 proteins; fold enrichment = 20.0; q = 7.98 × 10⁻³). Cell adhesion and signaling pathways were also enriched, including ‘MET activates PTK2 signaling’ (2 proteins; fold enrichment = 57.6; q = 3.23 × 10⁻³) and ‘Syndecan interactions’ (2 genes; fold enrichment = 48.9; q = 3.23 × 10⁻³). The proteins driving these enrichments included COL1A1, COL5A1, LUM, FBLN1, EFEMP1, and ICAM4, suggesting that dietary salt intake affects proteins involved in tissue structure and cell-matrix interactions. Although SVEP1 is known to be involved in extracellular matrix interactions, including integrin cell surface interactions, Reactome does not yet include SVEP1 in those pathways.

## Discussion

The major new finding of this study is that SVEP1 is upregulated in the plasma by high dietary sodium intake, and its upregulation correlates with less rise or a decrease in blood pressure during high-sodium diet compared with during low-sodium diet. Although SVEP1 was not the most significant protein in our analysis, it was highly significant, surviving false discovery rate correction and was internally validated by two aptamers. What drew our attention to SVEP1 is that it has been found to increase in other circumstances to increase in settings of volume expansion such as various forms of heart failure,^17-22^ but also pregnancy,^23^ in which volume increases and blood pressure decreases. Our data adds to the growing evidence that SVEP1 is involved in adaptation of the vascular system to increased blood volume, joining natriuretic peptides in such responses.

### SVEP1 as a Novel Mediator of Sodium Responses

The identification of SVEP1 as a prominent molecular response to dietary sodium represents a significant advance in understanding cardiovascular adaptation to sodium challenges. The prominence in our unbiased proteomic screen, confirmed by two independent aptamers and validated across different sodium states, establishes SVEP1 as a key mechanistic component in human sodium physiology. SVEP1 appears to be part of a coordinated response, suggesting that sodium adaptation involves tissue-level remodeling processes that extend beyond simple neurohumoral regulation, pointing toward structural adaptations that might support cardiovascular accommodation to sodium excess.

SVEP1’s prominence becomes more striking when compared to established biomarkers of sodium and volume regulation. An aptamer for NT-proBNP, the canonical marker of volume expansion, ranked only 16th (p=3.80×10⁻⁴) despite appropriate directional changes. Renin, a long-known sodium-regulatory hormone, ranked 23rd (p=7.33×10^⁻4^) with expected decline during high sodium intake. This hierarchy suggests that SVEP1 upregulation represents one of the body’s primary molecular responses to sodium loading.

We found that plasma SVEP1 and NT-proBNP levels were positively correlated overall (r = 0.69, p < 0.001), consistent with a prior report in heart failure cohorts in which SVEP1 showed a standardized coefficient of β = 0.54 with NT-proBNP.^17^ However, when we examined sodium-induced changes in these proteins, we observed a remarkably stronger correlation (r = 0.80, p < 0.001), with 64% of the variation in SVEP1 change explained by NT-proBNP change. This enhanced correlation likely reflects our controlled dietary intervention isolating their coordinated response to volume changes, whereas cross-sectional disease populations include heterogeneous factors—disease severity, medications, comorbidities—that obscure this relationship. The mean 1.4 kg weight change accompanying sodium loading and restriction support plasma volume as the shared regulatory stimulus driving both proteins.

This coordinated response suggests SVEP1 and NT-proBNP function as complementary biomarkers of volume status. SVEP1’s established prognostic value in hypertrophic cardiomyopathy,^18^ heart failure with reduced ejection fraction,^19^ and heart failure with supranormal ejection fraction^24^ might reflect its sensitivity to volume homeostasis. Notably, in hypertrophic cardiomyopathy, SVEP1 provides prognostic information independent of NT-proBNP,^18^ suggesting these proteins capture related but distinct aspects of cardiovascular adaptation to volume stress.

SVEP1’s known molecular functions provide plausible mechanisms for its action in adaptation to sodium excess. SVEP1’s sushi domains bind integrins α9β1 and α4β1 on vascular smooth muscle cells, triggering signaling pathways that reduce contractile force and promote vasodilation.^25^ This negative regulation of vasoconstriction could contribute to the vascular accommodation that prevents blood pressure elevation during volume expansion. Simultaneously, SVEP1’s C-terminal domains interact with Tie1 receptors on lymphatic endothelial cells, which promoted lymphangiogenesis and enhances fluid clearance capacity.^26-29^ Inhibition of lymphangiogenesis causes increased salt sensitivity in a rat model.^30^ A missense mutation in SVEP1 (p.D2702G) is associated with higher blood pressure.^31^ The allele associated with higher blood pressure is also associated with increased risk of coronary artery disease.^31^

### Convergent Evidence from Pregnancy Physiology

Independent proteomic analysis using the same SomaLogic platform during normal pregnancy revealed SVEP1 as the most upregulated plasma protein, increasing 24-fold during pregnancy.^23^ Chorionic villi transcriptomic analysis reveals a strong positive correlation between SVEP1 and gestational age (r = 0.7, p = 3.6 × 10^-5^).^32^ Single amino acid variant analysis suggests that at least some of this SVEP1 is supplied across the placenta to the mother.^33^ This convergent evidence from another physiological state of volume expansion paired with decreased blood pressure provides additional support for SVEP1’s role in cardiovascular adaptation to volume challenges.

### Study Limitations

Crossover studies have the limitation that carryover effects differing between the study arms can confound analysis of the effects of the treatment. We found no sequence effect with either SVEP1 aptamer (p>0.7 for each aptamer), indicating our washout period was of sufficient duration to allow permit valid analysis. Thus, carryover did not present a problem in this instance.

Our study was in a relatively small sample, although the crossover design and the strong contrast in dietary sodium amounts amplified the power to find effects of the dietary sodium. Those design features are likely why we found such strong statistical evidence of SVEP1’s potential role in vascular adaptation to high dietary sodium.

### Clinical Implications for Salt Sensitivity

The strong correlation between SVEP1 responses and blood pressure changes during sodium loading suggests potential clinical applications. SVEP1 measurement could serve as a biomarker for identifying individuals with robust adaptive capacity to sodium challenges versus those at risk for salt-sensitive hypertension.

The 2-fold higher SVEP1 responses in individuals with inverse salt sensitivity provide the first molecular signature for this phenotype. This finding could inform precision medicine approaches to sodium management, identifying individuals who might not benefit from aggressive sodium restriction.

### Therapeutic Implications

Understanding SVEP1’s role in sodium adaptation opens novel therapeutic directions. Interventions that enhance SVEP1 expression or activity could potentially improve sodium tolerance in salt-sensitive individuals. Given SVEP1’s dual functions in vascular relaxation and lymphangiogenesis, therapeutic approaches targeting these pathways may prove more effective than traditional volume management strategies.

The identification of coordinated extracellular matrix remodeling also suggests that sodium adaptation involves tissue-level changes that could be therapeutically modulated. The specific proteins identified in our analysis represent potential targets for enhancing cardiovascular accommodation to sodium challenges.

## Conclusions

Our findings challenge traditional approaches to understanding salt sensitivity that have focused primarily on renal mechanisms. The identification of SVEP1 and coordinated ECM responses suggests that cardiovascular adaptation to sodium involves complex changes to the vasculature that may be at least as important as renal regulation.

The convergent evidence from pregnancy--in which SVEP1 shows dramatic upregulation during another state of cardiovascular adaptation to increased plasma volume--suggests that SVEP1-mediated effects may represent fundamental mechanisms of cardiovascular accommodation that operate across multiple physiological contexts.

SVEP1 emerges as a prominent molecular correlate of blood pressure responses to dietary sodium in normotensive adults. The strong correlation between SVEP1 upregulation and blood pressure reduction during sodium loading, combined with SVEP1’s established functions in vascular and lymphatic biology, suggests that SVEP1-driven pathways mediate cardiovascular adaptation to sodium excess.

These findings provide the first molecular signature for inverse salt sensitivity and point toward precision medicine approaches for sodium management. The identification of coordinated extracellular matrix remodeling reveals the complexity of cardiovascular adaptation to sodium and suggests new therapeutic targets for managing salt sensitivity.

## Data Availability

The data will be made available in an appropriate repository once the manuscript is accepted for publication.

## Clinical Perspective

### What is new?

- SVEP1 emerges as a prominent molecular response to dietary sodium loading, ranking 4th among ∼7,000 proteins analyzed
- SVEP1 upregulation during high sodium intake correlates inversely with blood pressure changes, with 2-fold higher responses in inverse salt-sensitive individuals
- Coordinated extracellular matrix remodeling, including SVEP1, represents a novel pathway mediating cardiovascular adaptation to sodium challenges

### What are the clinical implications?

- SVEP1 measurement could serve as a biomarker to identify salt-sensitive versus salt-resistant individuals for personalized sodium management
- These findings may help identify the substantial subset of individuals (35% in this study) who experience blood pressure reduction with high sodium intake and may not benefit from sodium restriction
- SVEP1’s established roles in vascular relaxation and lymphangiogenesis suggest novel therapeutic targets for enhancing sodium tolerance in salt-sensitive individuals

## Acknowledgments

We thank the study participants and the University of Michigan’s Michigan Clinical Research Unit staff for their dedication to this research. We acknowledge SomaLogic Operating Co., Inc. as the provider of the proteomic data measured using the modified aptamer-based SomaScan® Assay. SomaScan®, SOMAmer® and SomaSignal™ are trademarks of SomaLogic Operating Co., Inc.

